# Accumulated Refusal Count: A Signal of Kidney Nonuse Risk (Brief Communication)

**DOI:** 10.64898/2026.01.08.26343720

**Authors:** Allan B. Massie, Leila Yan, Ruiqi Xue, Darren E. Stewart, Syed Ali Husain, Macey L. Levan, Sommer E. Gentry, Bonnie E. Lonze, Dorry L. Segev

**Affiliations:** Department of Surgery, New York University Grossman School of Medicine, New York, NY; Department of Medicine, New York University Grossman School of Medicine, New York, NY

**Author notes:** Contact Information: Allan B. Massie, Ph.D., Director, Quantitative Core, Center for Surgical and Transplant Applied Research, NYU Langone Transplant Institute, 1 Park Avenue, Sixth Floor, New York, NY 10016.

**Keywords:** kidney, nonuse, offer refusal count, allocation

## Abstract

A substantial proportion of recovered deceased-donor (DD) kidneys go unused. Accumulated refusals by transplant centers during the offer process may signal nonuse risk, and quantifying this phenomenon could inform frameworks for rescue strategies or out-of-sequence (OOS) placement. Using OPTN data on adult DD kidneys offered for transplant in 2024, we empirically estimated the probability of nonuse as a function of accumulated refusal count (ARC). Kidneys transplanted OOS were excluded from analysis. Among recovered adult DD kidneys offered in-sequence, risk of nonuse exceeded 50% after ARC=6 for blood type O kidneys, ARC=4 for type A and type B, and after ARC=1 for type AB. Risk exceeded 80% after ARC=128 (type O), ARC=55 (type A), ARC=50 (type B), and ARC=14 (type AB), and exceeded 90% after 980, 414, 278, and 41 refusals, respectively. The C-statistic of the ARC by blood type ranged from 0.896 to 0.933. ARC thresholds offer a pragmatic trigger for rescue allocation, incorporating center perception of kidney quality not easily captured in standard metrics. A policy allowing OPOs to offer kidneys OOS or deploy alternative rescue strategies once a certain ARC threshold is reached may improve utilization of hard-to-place donor kidneys while keeping easier-to-place kidneys in-sequence.

## 1. INTRODUCTION

A substantial proportion of deceased-donor (DD) kidneys recovered for transplant go unused. The nonuse rate has climbed over time, reaching 27.9% in 2023.^1^ Many factors may have contributed to this phenomenon, including a broadening of the DD organ pool,^2^ a labeling effect from the Kidney Donor Profile Index (KDPI),^3,4^ and the introduction of the KAS250 allocation system.^5^

Prospectively identifying DD kidneys at high risk of nonuse could inform frameworks for rescue allocation, out-of-sequence (OOS) placement, or related strategies to improve utilization and increase access to DD kidney transplantation (DDKT). Multifactorial risk scores have been proposed for this purpose,^6–10^ but these cannot account for nuanced clinical information such as anatomical abnormalities, abnormal machine perfusion values, or other detailed donor medical history that may contribute to difficulty in placing a DD kidney.^11^ As such, any risk score that relies entirely on clinical factors available for study in OPTN data will fail to accurately identify kidneys that are hard to place despite “looking good on paper”.

A simple, potentially useful signal to indicate risk of nonuse is the accumulating number of refusals as an OPO proceeds through a match run (“accumulated refusal count”, ARC). We hypothesized that increasing ARC would indicate higher risk of nonuse among DD kidneys. To test this hypothesis, we quantified the relationship between ARC and risk of nonuse among DD kidneys offered for KT January 2023-September 2025.

## 2. METHODS

### 2.1 Data source

The data reported in this study have been supplied by the United Network for Organ Sharing as the contractor for the Organ Procurement and Transplantation Network (OPTN). The OPTN data system includes data on all donors, waitlisted candidates, and transplant recipients in the United States, submitted by the members of the OPTN, and has been described elsewhere.^12^ This study used limited, patient-level Standard Transplant Analysis and Research files and offer data from July 2025. This study was determined not to constitute human subjects research by the NYU Langone Institutional Review Board.

### 2.2. Definition of ARC

For each DD kidney offered for transplantation in 2024, we counted the total number of offers of that kidney that were refused across all match runs. Multiple refusals of a donor kidney on behalf of the same patient in separate match runs were treated as separate events. We defined ARC as the (time-varying) number of accumulated refusals for a donor kidney. Thus, all kidneys started with ARC=0 when first recovered; after a single refused offer, a kidney’s ARC would equal 1; etc. Kidneys transplanted OOS were excluded from analysis.

### 2.3 Relationship between ARC and risk of nonuse

Using OPTN data on adult DD kidneys offered for DDKT in 2024, we empirically estimated the probability that a kidney would go unused as a function of ARC, stratified by blood type and calendar year. For example, within a blood type, probability of nonuse for ARC=10 equaled [number of DD kidneys with ≥10 refusals that went unused] / [total number of DD kidneys with ≥10 refusals]. All kidneys in the study population by definition had 0 or more refusals, and thus were used to calculate the nonuse rate for ARC=0. All kidneys refused at least once were used to calculate the nonuse rate for ARC=1, etc. To assess annual variation in the relationship between ARC and risk of nonuse, we then repeated the analysis for kidneys offered in 2023 and in January-September 2025.

### 2.4 Predictive validity of the ARC as a marker of nonuse risk

To quantify predictive validity of the ARC as a marker of nonuse risk, we calculated the C-statistic (area under the receiver operating curve), stratified by blood type and year. The C-statistic is calculated by comparing risk scores across each pair of observations that is discordant with respect to the outcome: here, each pair of kidneys in which one was transplanted and the other was not used. Since a kidney has no single ARC value but rather the ARC varies as refusals accumulate, for each such comparison we selected a random ARC for each organ between 0 and the total number of refusals for that kidney. For example, if a kidney was refused 12 times before being accepted, we selected a random ARC between 0 and 12; the ARC randomly selected for a given kidney varied from one comparison to the next.

## 3. RESULTS

### 3.1 Distribution of ORC

All donor kidneys started with an ORC of 0. Depending on ABO blood type, 51.8%-562.% of kidneys eventually reached ARC=10, 45.3%-48.3% reached ARC=20, and 36.2%-39.3% reached ARC=100 (Figure 1A). 33.3% of type O kidneys, 33.6% of type A kidneys, 31.3% of type B kidneys, and 9.3% of type AB kidneys reached ARC=400; 29.7% of type O kidneys, 27.7% of type A kidneys, 22.6% of type B kidneys, and 1.1% of type AB kidneys reached ARC=1000 (Figure 1B).

**Figure 1.**
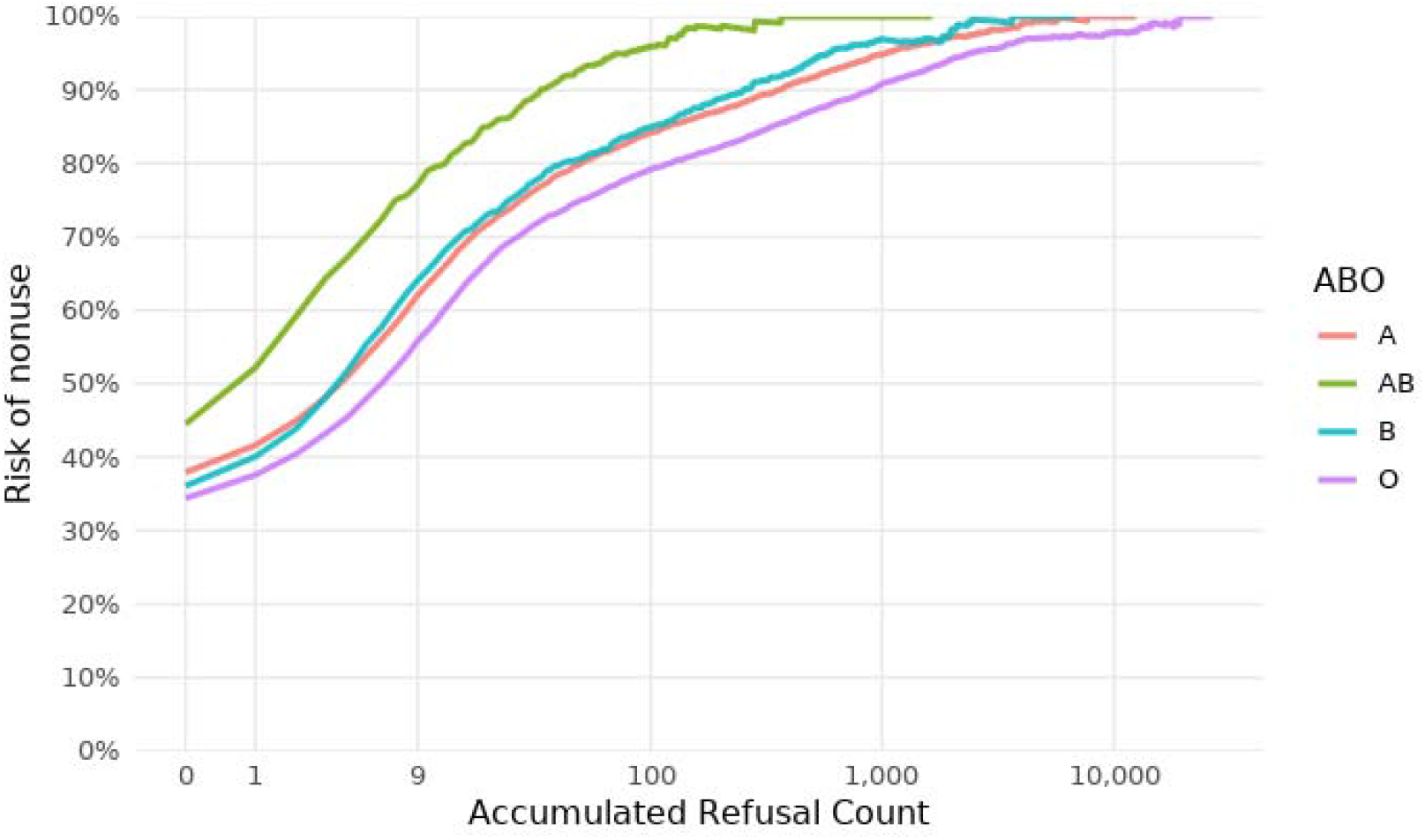
Risk of nonuse based on ARC, stratified by blood type. All kidneys in the study population by definition had 0 or more refusals, and thus were used to calculate the nonuse rate for ARC=0. All kidneys refused at least once were used to calculate the nonuse rate for ARC=1, etc. The X axis is presented on a logarithmic scale.

### 3.2 Relationship between the ARC and risk of nonuse

Among recovered adult deceased donor kidneys offered in-sequence in 2024, nonuse risk was 34.4% for type O kidneys, 36.1% for type B, 37.9% for type A, and 44.6% for type AB. Nonuse rates rose steadily as a function of ARC (Figure 1). The empirical risk of nonuse exceeded 50% after 6 accumulated refusals for type O kidneys, after 4 refusals for type A, after 4 for type B, and after 1 for type AB (Table 1). Risk exceeded 80% after 128 refusals (type O), 55 (type A), 50 (type B), and 14 refusals, respectively (type AB), and exceeded 90% after 980, 414, 278, and 41 refusals, respectively (Table 1).

**Table 1:** Thresholds of ARC at which risk of nonuse exceeds 50%, 80%, and 90%, stratified by blood type.

| **Blood type** | **>50% nonuse risk** | **>80% nonuse risk** | **>90% nonuse risk** |
| --- | --- | --- | --- |
| O | 6 | 128 | 980 |
| A | 4 | 55 | 414 |
| B | 4 | 50 | 278 |
| AB | 1 | 14 | 41 |

The thresholds that at which risk of nonuse exceeded 50%, 80%, and 90% were consistent across the three calendar years in the study period (Figure 2). Risk of nonuse exceeded 50% after fewer than 10 refusals for all ABO blood types in each year. Risk exceeded 80% after 120-150 refusals for type O kidneys, after 55-94 refusals for type A, after 50-74 refusals for type B, and after 14-23 refusals for type AB (Figure 2).

**Figure 2.**
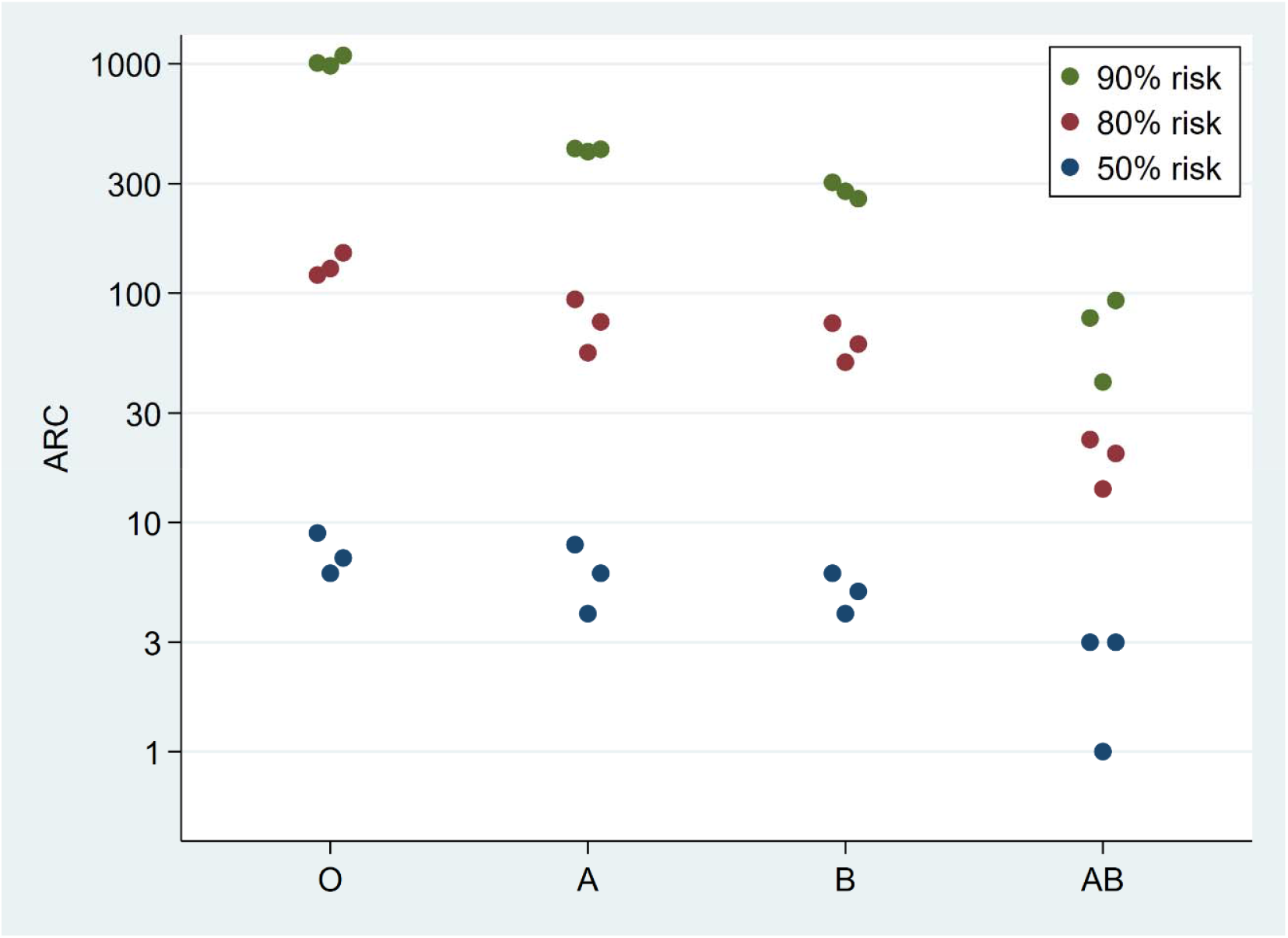
Thresholds of ARC at which risk of nonuse exceeds 50%, 80%, and 90%. Y axis is on a log scale. From left to right, the three dots represent 2023, 2024, and 1/2025-9/2025.

### 3.3 Predictive validity of the ARC as a marker of nonuse risk

The C-statistic of ARC ranged from 0.8963 to 0.9329, depending on calendar year and ABO blood type, indicating excellent predictive validity (Table 2). C-statistics were slightly lower for type AB (0.896-0.912), perhaps because match runs are shorter and there were more ties; C-statistics for the other three groups consistently exceeded 0.915 (Table 2).

**Table 2:** C-statistic of the ARC, by calendar year and blood type.

| **Blood type** | **2023** | **2024** | **2025** |
| --- | --- | --- | --- |
| O | 0.9244 | 0.9226 | 0.9183 |
| A | 0.9189 | 0.9203 | 0.9205 |
| B | 0.9176 | 0.9319 | 0.9329 |
| AB | 0.8998 | 0.9123 | 0.8963 |

## 4. DISCUSSION

In this national study of ARC and risk of nonuse, we found that risk of nonuse rose rapidly with accumulating refusals, exceeding 50% after fewer than ten refusals across all blood types, and 80% after a threshold ranging from 14 for type AB kidneys to 128 for type O kidneys in the year 2024; these thresholds were broadly consistent in 2023 and 2025. Predictive validity of the ARC as measured by the C-statistic ranged from 0.896 to 0.933, indicating excellent predictive validity.

Several multifactorial scores have previously been suggested for estimating risk of nonuse based on donor characteristics.^6–10^ C-statistics for these scores range from 0.83 to 0.89, indicating good predictive ability. However, none has been adopted for clinical use in the United States. The OPTN has used the KDPI, originally designed to predict donor-associated graft loss risk rather than nonuse, to facilitate accelerated placement.^13^ We have previously demonstrated that the KDPI predicts nonuse with a C-statistic of 0.82, inferior to indices designed for the purpose of predicting nonuse.^14^ The superior predictive validity of the ARC for predicting nonuse as compared to multifactorial indices likely results from the fact that ARC by nature incorporates all information that centers use in organ offer decisionOne barrier to adoption of a nonuse-based index might be the correct perception that such indices fail to capture all available information about a kidney. Just as some kidneys might be hard to place although they “look good on paper”, others (theoretically at least) could be relatively desirable despite a high risk score – “diamonds in the rough” that centers would want the opportunity to consider. A framework for rescue allocation or OOS placement based on ARC would allow at least some centers to evaluate an organ through a standard match run before switching to an alternative framework. Another drawback of a score based on donor characteristics is that the desirability of kidneys with certain donor characteristics might change over time. For example, several of the indices above identify donor hepatitis C as a marker of nonuse; however, the introduction of direct-acting antiviral medications has reduced risk to recipients of hepatitis C-positive kidneys, in turn increasing utilization of these kidneys.^15^ The ARC, based on center behavior, can adapt in real time to perceived desirability of a donor kidney.

Policies incorporating ARC as a trigger for OOS placement or related mechanism would incentivize OPOs to recover potentially hard-to-place kidneys, since they would feel confident that they could use every tool at their disposal to place the kidneys if standard allocation failed. Additionally, this framework would reduce offers of hard-to-place organs that are very unlikely to be accepted, reducing administrative burden on OPOs and centers and saving cold ischemia time, thereby likely further increasing utilization of hard-to-place organs. The risk threshold to trigger OOS placement or similar mechanism should be high enough that only genuinely hard-to-place kidneys are placed OOS, but low enough that the kidney is still viable. Ultimately, maximizing the number of deceased donor transplants benefits all patients by reducing the size of the waitlist.

Concern has been expressed that, in the process of implementing OOS to facilitate placement of challenging kidneys, some high-quality kidneys might also get placed OOS that could have easily been placed in-sequence.^16^ If OOS were allowed once a certain ARC trigger was reached under in-sequence allocation, this would not be a concern; desirable kidneys would be generally accepted in-sequence before the trigger was reached.

ARC thresholds offer a simple, pragmatic trigger for rescue allocation, incorporating OPO and center impression of kidney quality not easily captured in standard metrics. The ARC score has high predictive validity and its performance was consistent across the three years of our study period. A policy allowing OPOs to offer kidneys OOS or deploy alternative rescue strategies once a certain threshold of refusals is reached may improve utilization of hard-to-place donor kidneys.

## ABBREVIATIONS

DD: deceased donor
OOS: out of sequence
DDKT: deceased donor kidney transplantation
OPO: organ procurement organization
ARC: accumulated refusal count

## ACKNOWLEDGEMENTS

This work was supported by grant number R01DK132395 (Massie) from the National Institute of Diabetes and Digestive and Kidney Diseases (NIDDK). The analyses described here are the responsibility of the authors alone and do not necessarily reflect the views or policies of the Department of Health and Human Services, nor does mention of trade names, commercial products or organizations imply endorsement by the U.S. Government.

## DATA AVAILABILITY

The data used in this study are available from the Organ Procurement and Transplantation Network (OPTN).

